# Smoking during pregnancy and its effect on placental weight: A Mendelian randomization study

**DOI:** 10.1101/2023.08.24.23294537

**Authors:** Annika Jaitner, Marc Vaudel, Krasimira Tsaneva-Atanasova, Pål R. Njølstad, Bo Jacobsson, Jack Bowden, Stefan Johansson, Rachel M. Freathy

## Abstract

**Background:** The causal relationship between maternal smoking in pregnancy and reduced offspring birth weight is well established and is likely due to impaired placental function. However, observational studies have given conflicting results on the association between smoking and placental weight. We aimed to estimate the causal effect of newly pregnant mothers quitting smoking on their placental weight at the time of delivery.

**Methods:** We used one-sample Mendelian randomization, drawing data from the Avon Longitudinal Study of Parents and Children (ALSPAC) (up to N = 805) and the Norwegian Mother, Father and Child Cohort Study (MoBa) (up to N = 4475). The analysis was performed in pre-pregnancy smokers only, due to the specific role of the genetic instrument SNP rs1051730 (*CHRNA5 – CHRNA3 – CHRNB4*) in affecting smoking cessation but not initiation.

**Results:** Fixed effect meta-analysis showed a 175 g [95%CI: 16, 334] higher placental weight for pre-pregnancy smoking mothers who continued smoking at the beginning of pregnancy, compared with those who stopped smoking. Using the number of cigarettes smoked per day in the first trimester as the exposure, the causal estimate was a 12 g [95%CI: 2,22] higher placental weight per cigarette per day. Results were similar when the smoking exposures were measured at the end of pregnancy. Using the residuals of birth weight regressed on placental weight as the outcome, we showed weak evidence of lower offspring birth weight relative to the placental weight for continuing smoking.

**Conclusion:** Our results suggest that continued smoking during pregnancy causes higher placental weights.

**Key Messages:**

- It is well known that maternal smoking in pregnancy causes a lower birth weight on average, but the relationship between maternal smoking and placental weight is less clear, with observational studies showing conflicting results.
- Our Mendelian randomization study suggests that for pre-pregnancy smokers, continuing smoking during pregnancy causes higher placental weight at term than quitting smoking.
- Our study also suggests that a greater number of cigarettes smoked per day during pregnancy causes a larger placental weight at term.
- A possible explanation for our findings is that the placenta grows larger in mothers who smoke during pregnancy to compensate for the lower oxygen availability, but further work is needed to confirm and further investigate this hypothesis.

## Introduction

Maternal smoking during pregnancy is often described as one of the most modifiable risk factors for adverse pregnancy outcomes ^1^. Despite a strong public health message, many women continue to smoke in pregnancy. In the UK, the NHS digital service provides statistics indicating that approximately 8.6 % of mothers were known smokers at the time of delivery in the first half of 2023 ^2^. Mendelian Randomization studies between smoking during pregnancy and offspring birth weight suggest a causal relationship between smoking during pregnancy and lower birth weight ^3–6^. However, the underlying mechanisms remain unclear.

A potential mediator for the effect of smoking on fetal growth is the placenta, which provides oxygen and nutrient transport between mother and fetus ^7^. The maternal environment is experienced through the placenta ^8^. Additionally, studies have shown that maternal smoking is associated with altered histological morphology and structure, which, for example, can lead to a reduction in vascularization ^9,10^. Such abnormalities and the direct effect of nicotine on the placenta can reduce the maternal and fetal exchange, potentially leading to placental insufficiency ^11,12^. However, whether there is an effect of maternal smoking during pregnancy on placental weight is unclear because observational studies have yielded conflicting results. Wang et al.^7^ observed a reduction in placental weight in mothers who smoked in pregnancy versus non-smoking mothers. Furthermore, in a study of daily smokers who continued smoking throughout pregnancy, Larsen et al. ^13^ observed a linear decrease in placental weight with the number of cigarettes smoked per day for women who smoked throughout the whole of pregnancy. The same study also reported an increase in placental weight for women who stopped smoking after the first trimester compared to non-smokers ^13^. Similar results were reported by Mitsuda et al. ^14^ with the highest placental weights seen in women who quit smoking during pregnancy compared to never-smokers, those who gave up before pregnancy and those who smoked throughout pregnancy. But higher placental weights were seen in women who continued smoking in pregnancy compared to those who never smoked but also compared to those who quit before pregnancy ^14^. The apparently conflicting results from observational epidemiological studies linking smoking to placental weight may be due to unmeasured confounding and bias, and were conducted in different populations and with different study designs, making them difficult to compare. Hence, additional approaches are necessary to investigate a potential causal relationship.

We used one-sample Mendelian randomization to investigate any causal relationship between maternal smoking during pregnancy and placental weight in two studies: the Avon Longitudinal Study of Parents and Children (ALSPAC) ^15,16^ and the Norwegian Mother, Father and Child Cohort Study (MoBa) ^17,18^. Mendelian randomization (MR) enables the inference of causal effects in the presence of unobserved confounding through exploiting the natural randomization of inheritance of germline genetic variation from parents to their offspring happening at conception ^19^sup. We used a genetic variant, SNP rs1051730, as the instrumental variable to genetically proxy maternal smoking. Previous studies have shown that each additional copy of the risk allele rs1051730 is associated with higher odds of continuing smoking during pregnancy as well as an increase of about one cigarette per day ^20–22^. The SNP is located within the nicotine acetylcholine receptor gene cluster *CHRNA5 – CHRNA3 – CHRNB4* on chromosome 15. The biological relationship to smoking and nicotine dependence supports the association between the SNP and smoking. However, it is important to note that rs1051730 is not associated with smoking initiation ^20,22^. Due to the specific association of rs1051730 with smoking behaviour, we only used data from mothers who smoked before pregnancy to capture continuing smoking compared to stopping smoking in pregnancy. Figure 1 shows the directed acyclic graph describing the causal assumptions for our study analysis. Our aim was to improve the understanding of the effect of continuing smoking in pregnancy by investigating the causal relationship between maternal smoking and placental weight.

**Figure 1:**
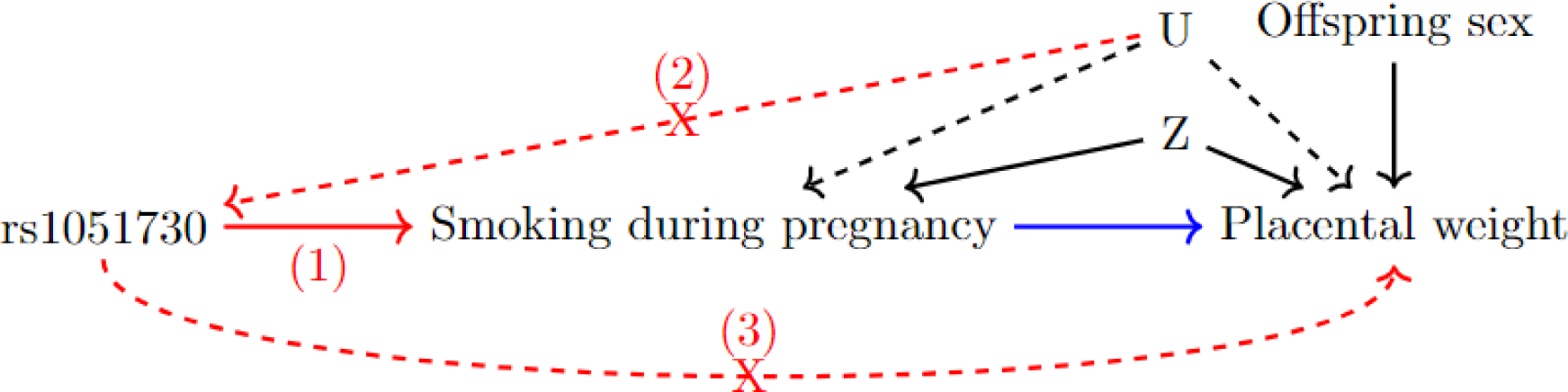
Directed acyclic graph (DAG) to highlight the Mendelian randomization (MR) framework. The MR assumptions for the instrumental variable (in this case maternal SNP rs1051730) are shown in red: 1. The instrumental variable needs to be associated with the exposure. 2. The instrumental variable is independent of confounding factors that confound the association of the exposure and the outcome. 3. The instrumental variable is independent of the outcome given the exposure and the confounding factors. The MR analysis estimates the effect between the exposure and the outcome shown in blue. The MR analysis is adjusted for offspring sex and ancestry principal components (and genetic batch variables in MoBa). These are summarised in the measured confounder variable Z. U stands for unmeasured confounders, which we are unable to include in the analysis.

## Methods

### Study populations

We performed our analysis in two different study populations. The **Avon Longitudinal Study of Parents and Children (ALSPAC)** is a prospective longitudinal cohort study ^15,16^. More information on the cohort is given in the supplementary material. We restricted our analysis to unrelated mothers with genetic information for the mother available. Additionally, we excluded multiple births and preterm births (pregnancy duration < 37 weeks). Full details including sample sizes are shown in Figure 2. Out of the unrelated mothers with genetic information available, about 37 % had recorded placental weight measures. After all exclusions, the analysis in pre-pregnancy smokers with available placental weight measures as an outcome was therefore performed in up to 805 individuals in ALSPAC. The **Norwegian Mother, Father and Child Cohort Study (MoBa)** is a population-based pregnancy cohort study conducted by the Norwegian Institute of Public Health ^17,18^. The study is linked with the **Medical Birth Registry of Norway (MBRN)**, a national health registry containing information about all births in Norway. More detailed information on the cohort and the version used is given in the supplementary material. We restricted the MoBa data to unrelated individuals with genetic information for the mother available. Additionally, we excluded multiple births and preterm births (pregnancy duration < 37 *7 days). Full details including sample sizes are shown in Figure 2. After all exclusions, the analysis in pre-pregnancy smokers with available placental weight measures as an outcome was performed in up to 4675 mothers in MoBa.

**Figure 2:**
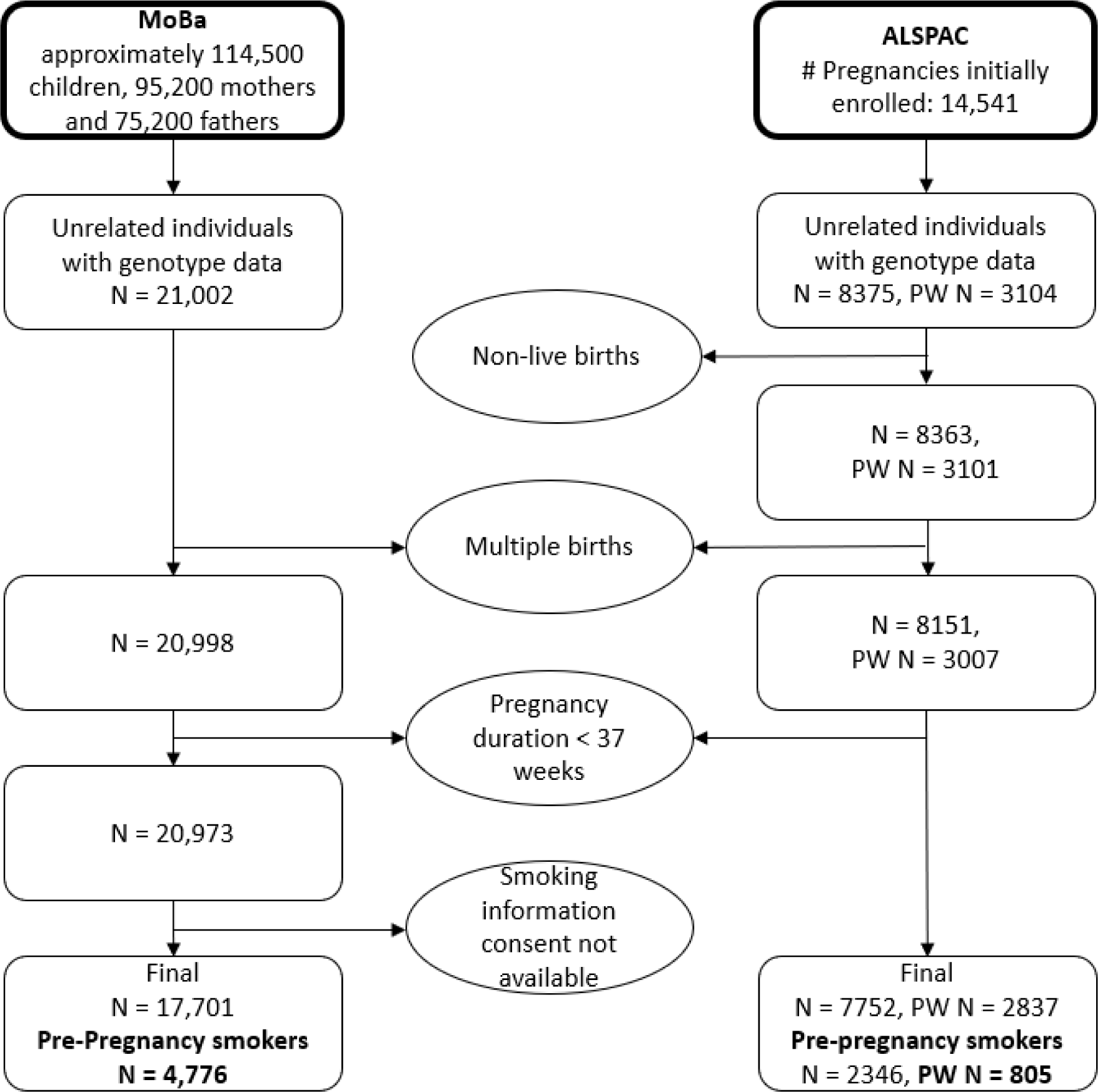
Flowchart to display the exclusion criteria of both the ALSPAC and the MoBa study. PW in the ALSPAC flow chart stands for placental weight and reflects the sample sizes with available placental weight measures. The sample sizes used for all Mendelian randomization analysis are highlighted in bold.

#### Genetic instrument

We instrumented the smoking behaviour using the single-nucleotide polymorphism (SNP) rs1051730, which has shown to be associated with smoking quantity and the inability to quit smoking but not smoking initiation ^20–22^. We used the genotype dosage of the genetic variant, rs1051730 as a continuous variable, which for each individual was a number close to 0, 1, or 2, reflecting the number of smoking risk alleles, combined with the probability of having 0, 1, or 2 risk alleles from the genotype imputation. More information on genotyping in both cohorts is described in detail in previously published articles^18,23^.

#### Outcome variable

The main outcome of interest is placental weight measured in grams. In ALSPAC, placental weight measures were obtained directly from obstetric records by research midwives who went back to the handwritten medical records of most patients and abstracted data including all weight measures. In MoBa, data related to pregnancy and birth were standardised and stem from the Medical Birth Registry of Norway (MBRN). Reporting placenta weight to the MBRN is mandatory and is carried out by the midwife attending the birth. All midwives share curriculum and training regarding the reporting of data, including examination of the afterbirth to the MBRN. The placenta is examined and characteristics of the placenta and umbilical cord, including measurements of the placental weight (untrimmed with the cord and membranes attached) are reported. The method has been unchanged since the inception of the MBRN in 1967. The reporting of these data to the MBRN has been validated, with good inter- and intra-observer agreement, making the data suitable for large scale epidemiological research ^24^.

#### Exposure variable

The exposures of interest were (i) continuing smoking during pregnancy vs. quitting and (ii) smoking quantity in cigarettes per day in the pre-pregnancy smokers. We used different measures of self-reported smoking variables. Study specific differences are outlined below.

#### Smoking variables of interest in ALSPAC

In ALSPAC, mothers were asked if they smoked before pregnancy. No specific time frame was given in the questionnaire to the mothers. We included everyone in the study who said they consumed tobacco before pregnancy even if this consumption was through other sources than cigarettes, such as pipes and cigars. The frequency of tobacco consumption via cigarettes was by far the highest (97.8 % of the mothers who smoked pre-pregnancy said they smoked cigarettes). The following smoking variables were used as exposures in the analysis performed in ALSPAC:

##### Smoking in the first three months of pregnancy

At 18 weeks of gestation the mother was asked whether she smoked in the first three months of pregnancy. This variable is self-reported and retrospective, however, any pregnancy complications post 18 weeks are not known at the time of the data collection.

##### Smoking in the last two weeks of pregnancy

This information was gained from a questionnaire sent out 8 weeks after the child was born. As for the previous variables smoking refers to any type of tobacco consumption. Note that for non-smoking mothers at this time point, the variable does not give information about whether the mothers stopped smoking at the beginning or throughout pregnancy.

##### Number of cigarettes smoked per day

Besides classifying whether a mother smoked or not as a binary variable, the participants were also asked about the number of cigarettes smoked per day at the same time points as previously described. The following categories were given: 0 cigarettes, 1-4, 5-9, 10-14, 15-19, 20-24, 25-29, 30 or more cigarettes. For the analysis the categories were coded with the number of the lower bound of each category.

#### Smoking variables of interest in MoBa

In MoBa, the mothers were asked whether they smoked during the last three months before becoming pregnant. The following smoking variables were used as exposures in the analysis performed in MoBa:

##### Mother smoking at the beginning of pregnancy

The information for this variable is from the MBRN. Data is preferably obtained from the antenatal health card which is filled out at the first antenatal visit between 6 and 12 weeks of gestation.

##### Mother smoking at the end of pregnancy

The information for this variable is from the MBRN. Data is preferably obtained from the antenatal health card. The end of pregnancy corresponds to the last trimester (approximately 36 weeks of gestation). As in ALSPAC, this variable does not contain information about the time point at which the mother stopped smoking and therefore is now classed as non-smoker.

##### Number of cigarettes smoked per day

The information for this variable is also taken from the Medical Birth Registry and reflects again the beginning and end of pregnancy separately. Different than in ALSPAC the number of cigarettes is not grouped into categories. The non-smokers with 0 cigarettes are initially not included. To allow comparison with ALSPAC, we coded the non-smokers as 0 based on the information given in the binary variables described above.

### Mendelian randomization

We performed one-sample Mendelian Randomization using individual level data. Mendelian randomization requires three assumptions to hold for rs1051730 to be a valid instrumental variable ^19^. The assumptions are graphically highlighted in Figure 1. Due to the genetic variants being defined at conception we assumed that it is independent of factors confounding the association between smoking during pregnancy and placental weight. We cannot formally test that the genetic instrument is only associated with the outcome through the exposure. However, based on the position of rs1051730 in the genome and therefore likely biological role, we assumed that the third assumption holds as well. Additionally, we studied the association between the SNP and various variables in the MoBa study and saw no associations (see supplementary SFigure 1).

For all analyses, we aimed to estimate the causal effect of smoking on placental weight (PW) in mothers who smoked pre-pregnancy (*S_pre_*=1). For continuous smoking definitions, our causal estimand was the population average effect of intervening to lower individuals observed smoking level *s* by 1 cigarette per day.

#### Continuous smoking

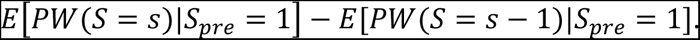

For binary smoking definitions, our causal estimand reflects the population average effect if all mothers continued to smoke versus if all mothers subsequently quit.

#### Binary smoking

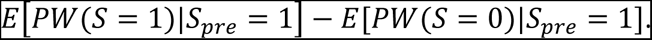

In each case, we impose a fourth identifying assumption of homogeneity, meaning that the causal effect does not vary across levels of a single instrument, nor across instruments. For all analysis a two stage regression approach was used. In the continuous smoking exposure case, the smoking variable (S) was firstly regressed on the SNP rs1051730 (G) and adjusted for known confounders or competing exposures (Z) via a linear model:

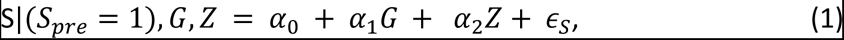

to furnish a genetically predicted smoking variable (^*S*^^). Secondly, PW was regressed on ^*S*^^ and known confounders or competing exposures (Z):

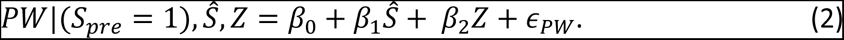

For binary smoking exposure variables, we performed a logistic regression in the first stage.

#### Residuals of z-score birth weight on z-score placental weight

We performed a final analysis by incorporating both birth and placental weight into a single outcome variable, thereby taking into account their relationship.

1) We firstly generated z-scores using generalised additive models for location, scale and shape from the gamlss R-package ^25,26^:
  a) of placental weight adjusting for gestational duration in female offspring (*PW*_*zf*_);
  b) of placental weight adjusting for gestational duration in male offspring (*PW*_*zm*_);
  c) of birth weight adjusting for gestational duration in female offspring (*BW*_*zf*f_);
  d) of birth weight adjusting for gestational duration in male offspring (*BW*_*zm*_).
  This resulted in adjusted z-scores of birth weight *BW*_*z*_ = (*BW*_*zm*_, *BW*_*zf*_) and placental weight *PW*_*z*_ = (*PW*_*zm*_, *PW*_*zf*_). The scores were derived from the individual level data within the ALSPAC and the MoBa study separately.
2) We then regressed *BW*_*z*_ on *PW*_*z*_:

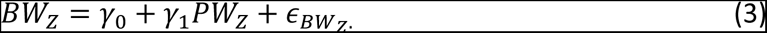
3) Next, we took the estimated residuals (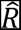) from the equation in step 2: 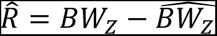.
4) Finally, we used the residuals from step 3 as the outcome in an MR analysis with a binary smoking exposure S, applying the two stage approach below:

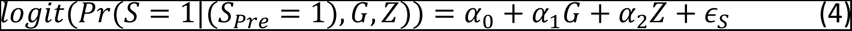

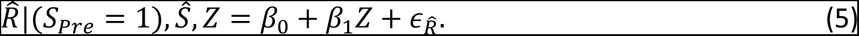 This enabled us to estimate the causal effect of maternal smoking on birth weight relative to placental weight.

#### Adjustment variables and meta-analysis analysis

We adjusted all analysis for offspring sex and principal components to account for population stratification (first 5 in ALSPAC and first 10 in MoBa). All analysis in MoBa were additionally adjusted for genetic batch variables. After performing the Mendelian randomization study in ALSPAC and in MoBa, we meta-analysed the results from smoking at the beginning of pregnancy and smoking at the end of pregnancy. The Q statistics (on 1df) (STable 1) provided no evidence to refute the null hypothesis that causal estimates derived from ALSPAC and MoBa pertained to different underlying quantities. We therefore combined them using an inverse variance weighted fixed effect model to produce an overall estimate.

## Results

### Study population characteristics

Table 1 shows clinical characteristics in the datasets used for the analysis from both the ALSPAC and the MoBa study.

**Table 1:** Descriptive statistics of the analysis data in the ALSPAC and the MoBa study. For the continuous variables mean and standard derivation are displayed. For the categorical variables a percentage is given. If there are two variable descriptions given in one row, then the first one corresponds to the ALSPAC study and the second one to the MoBa study. Not all variables are available in both dataset which results in missing values in the table.

| Variable | Mean (SD) / Percentage |  | N (no missing) |  |
| --- | --- | --- | --- | --- |
| Study | ALSPAC | MoBa | ALSPAC | MoBa |
| Birth weight (g) | 3402 (486) | 3674 (484) | 2346 | 4770 |
| Placental weight (g) | 654 (131) | 689 (145) | 804 | 4667 |
| Gestational duration (weeks in ALSPAC/days in MoBa) | 39.8 (1.3) | 282 (9) | 2346 | 4776 |
| Mothers age (years) | 26.8 (5) | 28.7 (4.7) | 2346 | 4776 |
| Mothers height (cm) | 164 (7) | 168.3 (5.8) | 2102 | 4735 |
| Mothers pre-pregnancy weight/<br>at the beginning of pregnancy (kg) | 62 (11) | 69 (13) | 2011 | 4660 |
| Mothers birth weight (g) | 3277 (631) | NA | 1289 | NA |
| Partners birth weight (g) | 3493 (768) | NA | 498 | NA |
| Fathers age at 15 weeks of gestation | NA | 31.3 (5.5) | NA | 4475 |
| Fathers height (cm) | NA | 181.6 (6.5) | NA | 4696 |
| Fathers weight (kg) | NA | 85.9 (12.7) | NA | 4647 |
| Offspring (male %) | 50.8 | 50.3 | 2346 | 4776 |
| Smoking in the first three month of<br>pregnancy/ Smoking at the beginning<br>of pregnancy (yes %) | 72.4 | 35 | 2346 | 4715 |
| Smoking in the last two weeks of<br>pregnancy/Smoking at the end of<br>pregnancy (yes %) | 57.9 | 17.5 | 2032 | 4580 |
| Year of delivery | 1991-1993 | 1999-2009 | 2346 | 4776 |

### SNP-exposure association in ALSPAC and MoBa

The results for the association between the different smoking exposures and the genetic instrument rs1051730 are shown in Table 2. This corresponds to the first stage of the Mendelian randomization. For all the different smoking variables the SNP is a strong instrument showing that each additional risk allele increases the likelihood of continuing smoking in pregnancy as well as the quantity.

**Table 2:** Effect estimates of the smoking variables regressed on the genetic instrument as shown in equation (1). The analysis is adjusted for offspring sex and the principal components (and genetic batch variables in MoBa). For the first 4 rows a logistic regression is used due to the smoking variables being binary. The effect estimate in the first column gives the odds of continuing smoking in pregnancy per additional risk allele of rs1051730. For the last 4 rows a linear regression is used. The effect estimate in the first column gives the change in the number of cigarettes smoked per day per each additional risk allele of rs1051730.

| Smoking variable | Effect Estimate | Lower 95 % CI | Higher 95 % CI | P-value | F-statistic | N | Study |
| --- | --- | --- | --- | --- | --- | --- | --- |
| Smoking in the first three months of pregnancy | 1.27 | 1.11 | 1.46 | 0.0009 | 11.1 | 804 | ALSPAC |
| Smoking in the last two weeks of pregnancy | 1.25 | 1.09 | 1.43 | 0.0015 | 10 | 691 | ALSPAC |
| Smoking at the beginning of pregnancy | 1.15 | 1.04 | 1.27 | 0.0016 | 9.9 | 4606 | MoBa |
| Smoking at the end of pregnancy | 1.27 | 1.13 | 1.43 | 0.00004 | 17.1 | 4475 | MoBa |
| # of cigarettes smoked per day in the first three month | 0.88 | 0.23 | 1.338 | 0.0001 | 14.7 | 790 | ALSPAC |
| # of cigarettes smoked per day in the last two weeks | 0.87 | 0.25 | 1.36 | 0.0004 | 12.7 | 690 | ALSPAC |
| # of cigarettes smoked per day at the beginning | 0.45 | 0.11 | 0.67 | 0.00003 | 17.2 | 4267 | MoBa |
| # of cigarettes smoked per day at the end | 0.25 | 0.06 | 0.37 | 0.00006 | 16 | 4341 | MoBa |

### Binary smoking exposure

In the fixed effect meta-analysis, we observed that mothers who continued smoking in pregnancy had, on average, a 175 g (95% CI: [16,334]) higher placental weight compared with those who stopped smoking at the beginning of pregnancy. The F-statistic as a measure of the strength of the instrument was 11.1 in ALSPAC and 9.9 in MoBa for the analysis at the beginning of pregnancy, which is very close to the minimum F-Statistic of 10 suggested in the literature ^19,27^. In MoBa, the F-Statistic, 17.1, was higher for the analysis with the smoking at the end of pregnancy exposure. In ALSPAC, the F-Statistic did not change much for the different time points of smoking in pregnancy as the exposure. For both ALSPAC and MoBa, and the meta-analysis similar effect sizes were evident for the analysis at the end of pregnancy (meta-analysis: 193 g, 95% CI: [40,346]) compared to the analysis with the smoking exposure being measured at the beginning of pregnancy. Results for the Mendelian randomization study in pre-pregnancy smokers in ALSPAC and in MoBa as well as the fixed effect meta-analysis for a binary smoking exposure are displayed in Figure 3.

**Figure 3:**
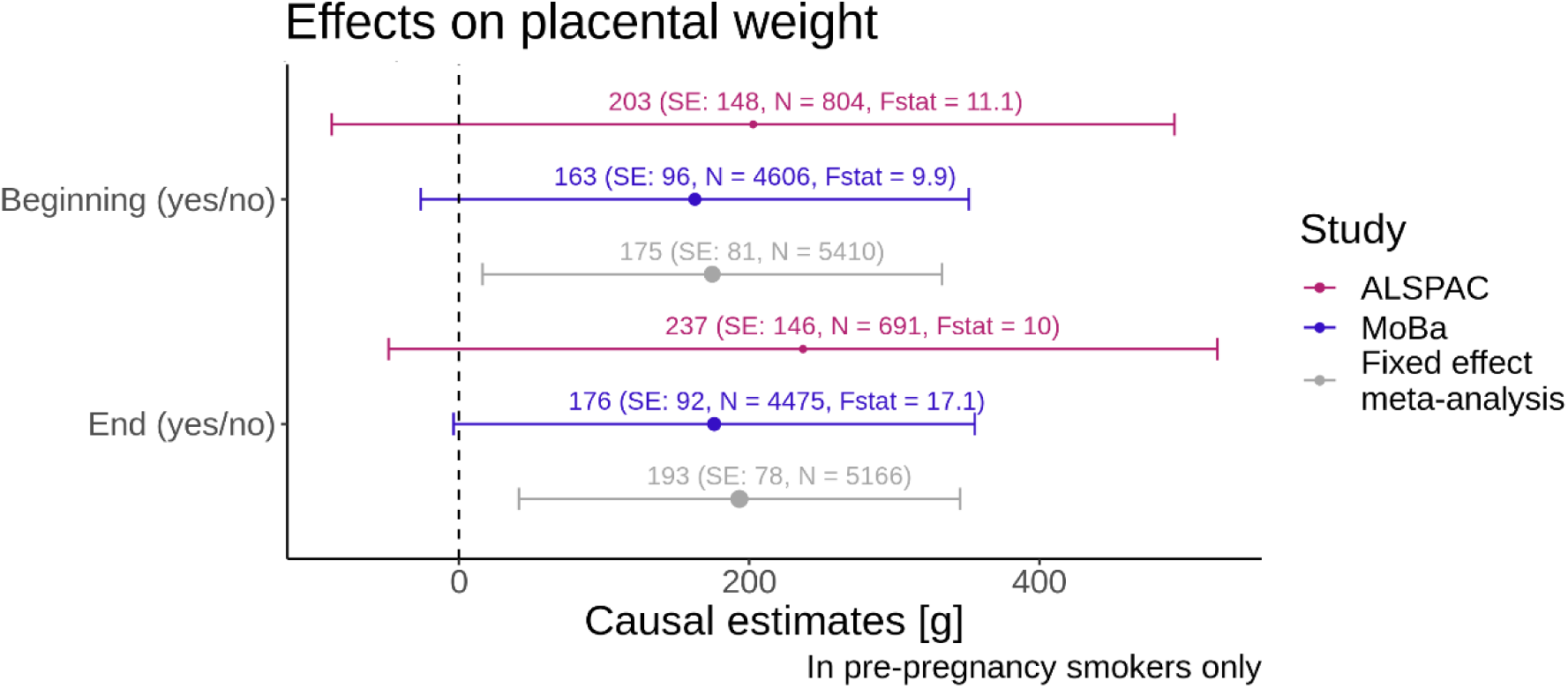
Forest plot with binary smoking variables on the y-axis and the causal estimate from the Mendelian randomization with placental weight as the outcome on the x-axis. The colours indicate the results for the different studies and the fixed effect meta-analysis. The bars indicate the 95% confidence intervals. The F-statistics from the first stage of the MR analysis are displayed alongside with the sample size N for each analysis. The size of the dot of the point estimate for each analysis is proportional to 1/SE.

### Cigarettes smoked per day exposure

The meta-analysis results indicated an increase of 12 g (95% CI: [2,22]) in placental weight for each additional cigarette smoked at the beginning of pregnancy amongst the mothers who smoked before pregnancy. Each additional cigarette at the end of pregnancy caused an increase in placental weight of 18 g (95% CI: [4,32]). Figure 4 shows the results of this Mendelian randomization study. The effects in ALSPAC and MoBa were consistent with the meta-analysis. For these analyses the F-statistics were slightly higher than for the binary analysis.

**Figure 4:**
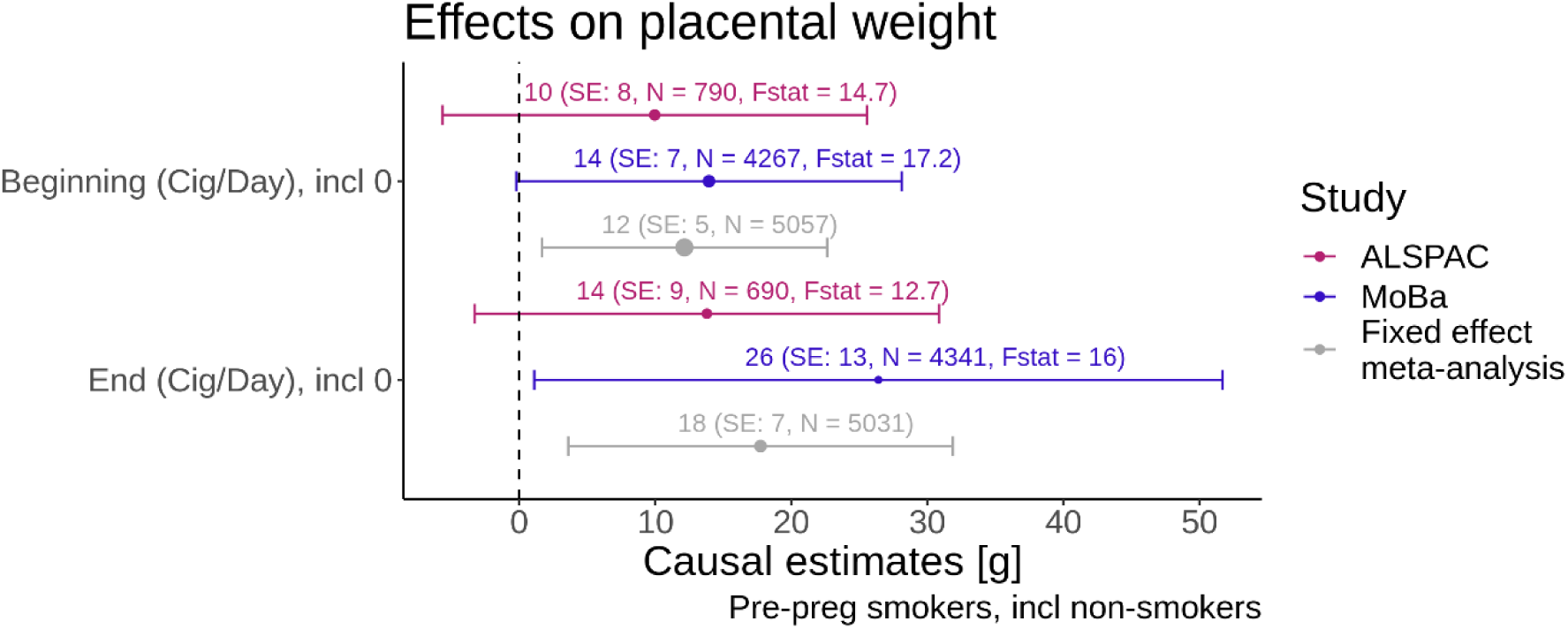
Forest plot with smoking quantity variables on the y-axis and the causal estimate from the Mendelian randomization with placental weight as the outcome on the x-axis. The colours indicate the results for the different studies and the fixed effect meta-analysis. The bars indicate the 95% confidence intervals. The F-statistics from the first stage of the MR analysis are displayed alongside with the sample size N for each analysis. The size of the dot of the point estimate for each analysis is proportional to 1/SE.

**Figure 5:**
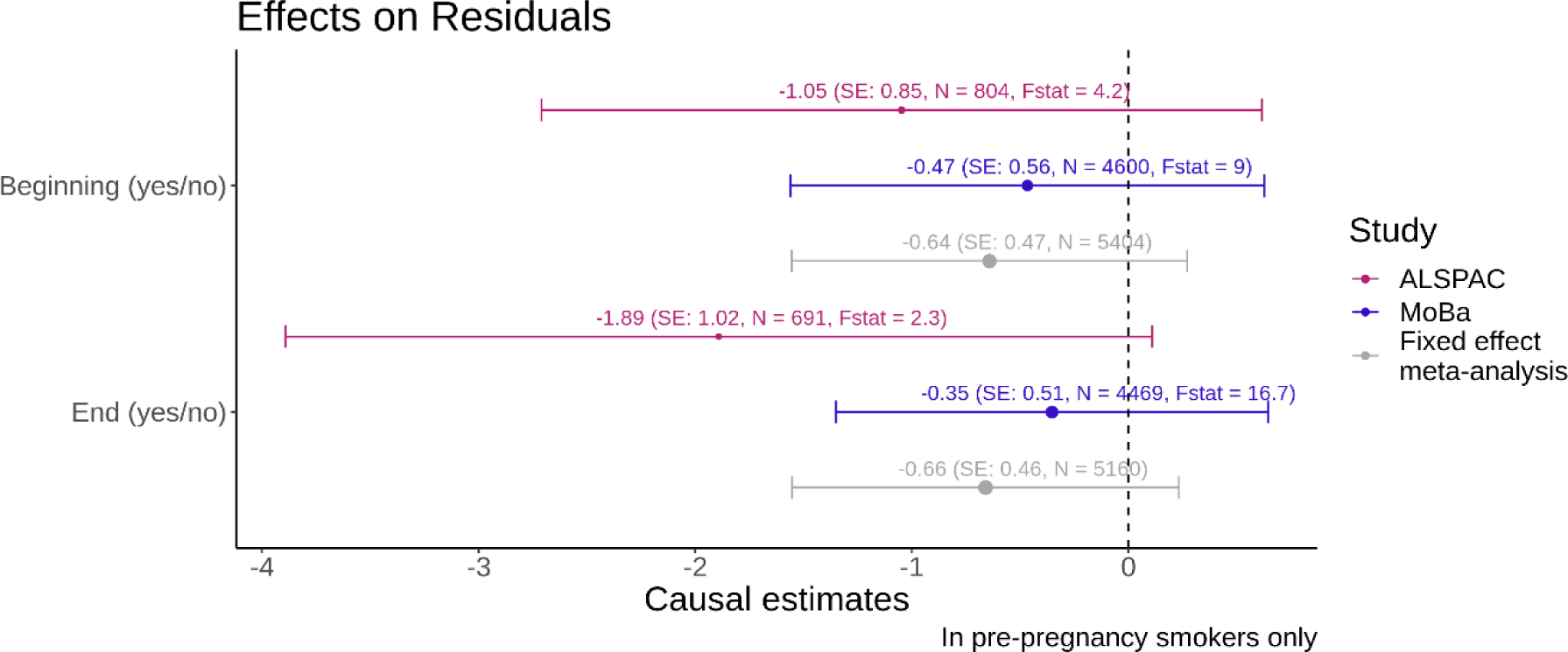
Forest plot with binary smoking variables on the y-axis and the causal estimate from the Mendelian randomization with the residuals of the regression of adjusted z-score birth weight on adjusted z-score on placental weight as the outcome on the x-axis. The effect sizes reflect the change in z-score birth weight adjusted for z-score placental weight for continuing smoking vs quitting in pregnancy. The colours indicate the results for the different studies and the fixed effect meta-analysis. The bars indicate the 95% confidence intervals. The F-statistics from the first stage of the MR analysis are displayed alongside with the sample size N for each analysis. The size of the dot of the point estimate for each analysis is proportional to 1/SE.

### Residual z score analysis

For both ALSPAC and MoBa, negative point estimates were obtained for the Mendelian Randomization with the residuals of the regression of adjusted birth weight on adjusted placental weight as outcome. This indicated that for mothers who continue to smoke, their offspring birth weights tend to be lower relative to the placental weight. However, although the results were consistent, it should be noted that 95% confidence intervals for both the individual study estimates, and the meta-results overlap with zero.

## Discussion

We have shown that continuing smoking during pregnancy is causally associated with higher placental weights using a Mendelian randomization approach. The results were consistent for both the binary exposure of continuing vs quitting smoking and number of cigarettes smoked per day in two independent cohorts.

Given the well-established relationship between maternal smoking in pregnancy and lower birth weight, it is plausible that smoking would lead to lower placental weights due to an impairment of the placental function. Zdravkovic et al.^28^ stated the likelihood of tobacco exposure reducing the blood flow between mother and child thereby causing a hypoxic environment for the fetus and this could be manifested in decreased placental and fetal growth as oxygen binding is essential for the development of these organs. However, our findings are more in line with a compensatory effect. The placenta might grow larger relative to birth weight to meet the oxygen demands of the fetus and to restore oxygen binding sites. This hypothesis is supported by our findings of the residual analysis, which showed lower birth weights relative to the placental weight for mothers who continue to smoke vs those who quit smoking in pregnancy. The impact of a hypoxic environment on the placenta has been studied in animal models with conflicting results ^29^. For example, increased placental weights with a reduced fetal weight were seen in guinea pigs when exposed to chronic mid gestation 10.5 % hypoxia ^30^ and observed in mice for a chronic early 13 % hypoxia ^31^. Studies in rats have reported that under chronic 13-14 % hypoxia in early gestation, an increased placental weight was detected, but without any change in fetal weight ^32,33^. This suggests that in some conditions the placenta might be able to adapt and compensate for the hypoxic environment, but in other situations, the enhanced placental growth (and therefore placental weight increase) limits other factors of the fetal development process. Placental weight is often used to proxy placental function ^34^, but as discussed above this is not a straightforward relationship and needs to be carefully interpreted. Additionally, placenta weight is a combination of several components including size, surface area and thickness. Both abnormally higher and lower placental weights are associated with increased risk of pregnancy complications ^29^. Further explorations of the placenta, placenta functioning and efficacy and how to quantify these are necessary. However, sample sizes of such studies are currently limited and a Mendelian randomization study to investigate causality was not feasible.

Due to the properties of the Mendelian randomization method, adjusting for confounders is not strictly necessary but can increase precision. However, it is important to only adjust for variables that, one is confident about, act as a confounder to the exposure and the outcome variable. Therefore, we adjusted all our analysis for offspring sex and the population stratification via principal components. There are various other covariates that we could have adjusted for, like, for example, gestational duration. However, it is possible that gestational duration acts as a mediator for smoking in pregnancy and placental weight rather than a confounder. Adjusting for gestational duration could then induce collider bias. In the supplementary material (SFigure 3) we showed that additionally adjusting the MR analysis for different sets of covariates, which are potential confounders of the relationship between the smoking exposure and placental weight, were consistent with the results in the main paper.

One of the limitations of our study is that the available sample size of mothers who smoke at the beginning of pregnancy was limited. Hence, this leads to large uncertainties surrounding the magnitude of the effect on placental weight. However, this study comprises two of the biggest mother child birth cohorts available and the results across all the different analysis models in both cohorts are consistent. Another limitation is that all smoking information from the mothers was self-reported data. The strong public health message on smoking might potentially lead to underreporting of smoking in pregnancy. However, a validation of self-reported smoking was performed in a subset of the MoBa participants and revealed that daily smoking prevalence increased only slightly, from 9% to 11%, when investigating cotinine concentrations, suggesting that self-reported smoking is a valid marker for tobacco exposure in MoBa ^35^.

One of the strengths of our study is the use of the SNP rs1051730 which has very robust statistical evidence for association with smoking cessation and smoking quantity. There is also strong biological evidence for this association as SNP rs1051730 is in the nicotine acetylcholine receptor gene cluster *CHRNA5-CHRNA3-CHRNB4.* Rare variant burden associations have implicated all three of these genes as important in influencing smoking quantity ^36^.

In conclusion, this study suggests that maternal smoking leads to a compensatory increase in placenta weight, but further investigations on maternal smoking, birth weight and placental properties are necessary to better understand mediation effects or other forms of interactions between these three components.

## Ethics approval

The establishment of MoBa and initial data collection was based on a licence from the Norwegian Data Protection Agency and approval from The Regional Committees for Medical and Health Research Ethics. The MoBa cohort is currently regulated by the Norwegian Health Registry Act. The current study was approved by the Regional Committees for Medical and Health Research Ethics (no. 2012/67).

Ethical approval for the study was obtained from the ALSPAC Ethics and Law Committee and the Local Research Ethics Committees. Informed consent for the use of data collected via questionnaires and clinics was obtained from participants following the recommendations of the ALSPAC Ethics and Law Committee at the time.

## Data Availability

All information on data availability is decribed in detail in the manuscript

## Data Availability

The data in ALSPAC is fully available, via managed systems, to any researchers. The managed system is a requirement of the study funders, but access is not restricted on the basis of overlap with other applications to use the data or on the basis of peer review of the proposed science.

The ALSPAC data management plan describes in detail the policy regarding data sharing, which is through a system of managed open access. The following steps highlight how to apply for access to the data included in this paper and all other ALSPAC data. (1) Please read the ALSPAC access policy, which describes the process of accessing the data and samples in detail and outlines the costs associated with doing so. (2) You may also find it useful to browse the fully searchable ALSPAC research proposals database, which lists all research projects that have been approved since April 2011. (3) Please submit your research proposal for consideration by the ALSPAC Executive Committee. You will receive a response within 10 working days to advise you whether your proposal has been approved. If you have any questions about accessing data, please.

Data from the Norwegian Mother, Father and Child Cohort Study and the Medical Birth Registry of Norway used in this study are managed by the national health register holders in Norway (Norwegian Institute of public health) and can be made available to researchers, provided approval from the Regional Committees for Medical and Health Research Ethics (REC), compliance with the EU General Data Protection Regulation (GDPR) and approval from the data owners. The consent given by the participants does not open for storage of data on an individual level in repositories or journals. Researchers who want access to data sets for replication should apply through helsedata.no. Access to data sets requires approval from The Regional Committee for Medical and Health Research Ethics in Norway and an agreement with MoBa.

## Supplementary data

Supplementary data are available online.

## Author contributions

A.J., R.M.F., J.B. and K.T.A, contributed to the study design. A.J. performed the analyses in this study and drafted the manuscript. Data interpretation and statistical analysis were aided by J.B. and K.T.A.. Biological and clinical interpretation were supported by R.M.F., S.J. and B.J.. P.R.N., S.J. and M.V. contributed to the collection of and management of the MoBa cohort data. For the analysis in the MoBa dataset A.J. was supported by M.V. and S.J. All authors reviewed and edited previous versions of the manuscript. All authors read and approved the final manuscript.

## Funding

The UK Medical Research Council and Wellcome (Grant ref: 217065/Z/19/Z) and the University of Bristol provide core support for ALSPAC. This publication is the work of the authors and A.J. and R.M.F. will serve as guarantors for the contents of this paper.

Genotyping of the ALSPAC maternal samples were funded by the Wellcome Trust (WT088806). Specific funds for recent detailed data collection on the mothers were obtained from the US National Institutes of Health (R01 DK077659) and Wellcome Trust (WT087997MA) for completion of selected items of obstetric data extraction, including placental weights. A comprehensive list of grants funding is available on the ALSPAC website (http://www.bristol.ac.uk/alspac/external/documents/grant-acknowledgements.pdf).

A.J. received funding for her PhD studentship from the Faculty of Health and Life Sciences at the University of Exeter

P.R.N. was supported by grants from the European Research Council (AdG #293574), Trond Mohn Foundation (Mohn Center for Diabetes Precision Medicine), the Research Council of Norway (#240413), the Western Norway Regional Health Authority (Strategic Fund), the Novo Nordisk Foundation (#NNF54741).

K.T.A. gratefully acknowledges the financial support of the EPSRC via grant EP/T017856/1.

S.J. was supported by Helse Vest’s Open Research Grant (grants #912250 and F-12144), the Novo Nordisk Foundation (NNF20OC0063872) and the Research Council of Norway (grant #315599).

M.V. acknowledges the support of the Research Council of Norway (project #301178)

R.M.F. is supported by a Wellcome Senior Research Fellowship (WT220390).

This project utilised high-performance computing funded by the UK Medical Research Council (MRC) Clinical Research Infrastructure Initiative (award number MR/M008924/1).

This study was supported by the National Institute for Health and Care Research Exeter Biomedical Research Centre. The views expressed are those of the authors and not necessarily those of the NIHR or the Department of Health and Social Care.

This research was funded in part, by the Wellcome Trust (Grant number: WT220390). For the purpose of Open Access, the author has applied a CC BY public copyright licence to any Author Accepted Manuscript version arising from this submission.

## Acknowledgements

We are extremely grateful to all the families who took part in this study, the midwives for their help in recruiting them, and the whole ALSPAC team, which includes interviewers, computer and laboratory technicians, clerical workers, research scientists, volunteers, managers, receptionists and nurses.

The Norwegian Mother, Father and Child Cohort Study is supported by the Norwegian Ministry of Health and Care Services and the Ministry of Education and Research. We are grateful to all the participating families in Norway who take part in this on-going cohort study.

We thank the Norwegian Institute of Public Health (NIPH) for generating high-quality genomic data. This research is part of the HARVEST collaboration, supported by the Research Council of Norway (#229624). We also thank the NORMENT Centre for providing genotype data, funded by the Research Council of Norway (#223273), South East Norway Health Authorities and Stiftelsen Kristian Gerhard Jebsen. We further thank the Center for Diabetes Research, the University of Bergen for providing genotype data and performing quality control and imputation of the data funded by the ERC AdG project SELECTionPREDISPOSED, Stiftelsen Kristian Gerhard Jebsen, Trond Mohn Foundation, the Research Council of Norway, the Novo Nordisk Foundation, the University of Bergen, and the Western Norway Health Authorities.

Supported by grants from the European Research Council (AdG #293574), the Bergen Research Foundation (“Utilizing the Mother and Child Cohort and the Medical Birth Registry for Better Health”), Stiftelsen Kristian Gerhard Jebsen (Translational Medical Center), the University of Bergen, the Research Council of Norway (FRIPRO grant #240413), the Western Norway Regional Health Authority (Strategic Fund “Personalized Medicine for Children and Adults”), the Novo Nordisk Foundation (grant #54741), and the Norwegian Diabetes Association; and (to S.J.) Helse Vest’s Open Research Grant (grant #912250), the Research Council of Norway (FRIPRO grant #315599), and Novo Nordisk Foundation (grant #NNF21OC0070349). This work was partly supported by the Research Council of Norway through its Centres of Excellence funding scheme (#262700), Better Health by Harvesting Biobanks (#229624) and The Swedish Research Council, Stockholm, Sweden (2015-02559), The Research Council of Norway, Oslo, Norway (FRIMEDBIO #547711), March of Dimes (#21-FY16-121). The Norwegian Mother, Father, and Child Cohort Study is supported by the Norwegian Ministry of Health and Care Services and the Ministry of Education and Research, NIH/NIEHS (contract no N01-ES-75558), NIH/NINDS (grant no.1 UO1 NS 047537-01 and grant no.2 UO1 NS 047537-06A1).

Analyses were performed using digital laboratories in HUNT Cloud at the Norwegian University of Science and Technology, Trondheim, Norway. We are grateful for outstanding support from the HUNT Cloud community.

The authors would like to acknowledge the use of the University of Exeter High-Performance Computing (HPC) facility in carrying out this work.

