## Supplementary material for "Smoking during pregnancy and its effect on placental weight: A Mendelian randomization study"

### Study population information

The **Avon Longitudinal Study of Parents and Children (ALSPAC)** is a prospective longitudinal cohort study. Pregnant women resident in Avon, UK with expected dates of delivery between 1st April 1991 and 31st December 1992 were invited to take part in the study. A total of 20,248 pregnancies were identified as being eligible and the initial number of pregnancies enrolled was 14,541. Of the initial pregnancies, there was a total of 14,676 foetuses, resulting in 14,062 live births and 13,988 children who were alive at 1 year of age.

The **Norwegian Mother, Father and Child Cohort Study (MoBa)** is a population-based pregnancy cohort study conducted by the Norwegian Institute of Public Health. Participants were recruited from all over Norway from 1999-2008. The women consented to participation in 41% of the pregnancies. The cohort includes approximately 114,500 children, 95,200 mothers and 75,200 fathers. The current study is based on version 10 of the quality-assured data files released for research in 2018_02_01.
The **Medical Birth Registry (MBRN)** is a national health registry containing information about all births in Norway.

### The Q statistics (on 1df) provided no evidence to refute the null hypothesis that causal estimates derived from ALSPAC and MoBa pertained to different underlying quantities.

| Exposure | Q-statistic | P-value |
|---|---|---|
| Beginning (yes/no) | 0.05 | 0.82 |
| End (yes/no) | 0.13 | 0.72 |
| Beginning (Cig/Day), incl 0 | 0.14 | 0.71 |
| End (Cig/Day), incl 0 | 0.66 | 0.42 |
| Residual z-score analysis: Beginning (yes/no) | 0.33 | 0.57 |
| Residual z-score analysis: End (yes/no) | 1.82 | 0.18 |

*STable 1: Q-statistic results and p-values to test for heterogeneity in the fixed effect meta-analysis. First column refers to the exposures used in each meta-analysis. The first two rows correspond to the binary exposure analysis, the third and fourth row corresponds to the meta-analysis with the number of cigarettes smoked per day , the last two rows correspond to the meta-analysis with the residual z-scores as outcome and a binary smoking exposure. All results suggest the absence of heterogeneity.*

### Sensitivity analysis: The SNP is not associated with a variety of variables.

One of the assumptions for a valid instrumental variable is that the instrument is only associated with the outcome through the exposure of interest. This means we assume no pleiotropic effect. The assumption cannot be tested directly. However, we performed a sensitivity analysis showing that the SNP is not associated with a set of variables that could affect placental weight and therefore are a potential for pleiotropic pathways. Based on the location of the SNP rs1051730, which we used as instrumental variable in the MR analysis, we did not expect any associations and SFigure 1 confirms that there are no associations between the SNP and a set of different variables.

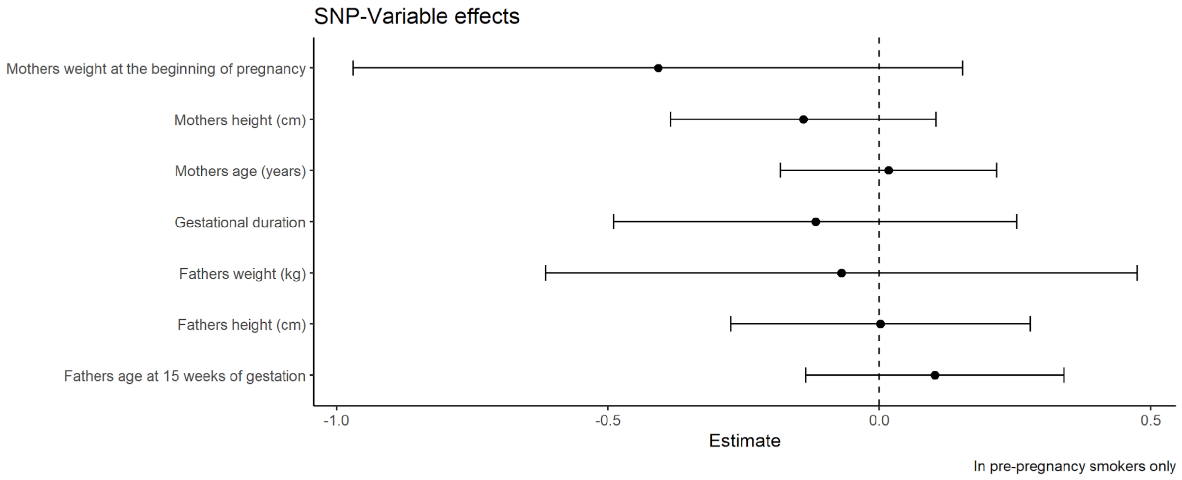


*SFigure 1: Results for the association between the SNP rs1051730 and a set of different variables shows no associations for all the analysis. We only used data from the MoBa study and as in the rest of this paper restricted the analysis to mothers who smoked before pregnancy.*

### Sensitivity analysis: Adjusting the MR analysis for different covariates is consistent with the results in the main paper of continuing smoking during pregnancy causing higher placental weights.

The MR analysis, in the main part of the paper, investigating the effect of continuing smoking in pregnancy on placental weight was only adjusted for offspring sex. We believe this is a competing exposure and therefore adjusting for it will increase the precision of the analysis. In addition, we looked at adjusting for different covariate sets including combinations of the variables: offspring sex, gestational duration, paternal smoking before pregnancy and paternal smoking at 15 weeks of gestation. Note that the information about the smoking habits of the father of the baby were obtained through the questionnaire for the mother. The results of adjusting for different covariates are given in SFigure 3 and the DAG for this analysis is given in SFigure 2, where Z represents the different covariates for which we adjusted in this sensitivity analysis. We show that including paternal smoking information into the confounder set reduced the point estimate. However, all the estimates are consistent with the results in the main part of this paper showing an increased placental weight for mothers who continue to smoke in pregnancy.

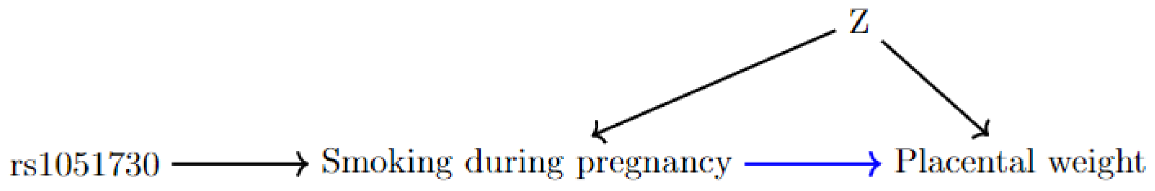


*SFigure 2: DAG for the sensitivity analysis of adjusting the MR analysis for different covariates. Z varies and represents the different covariates we adjusted for.*

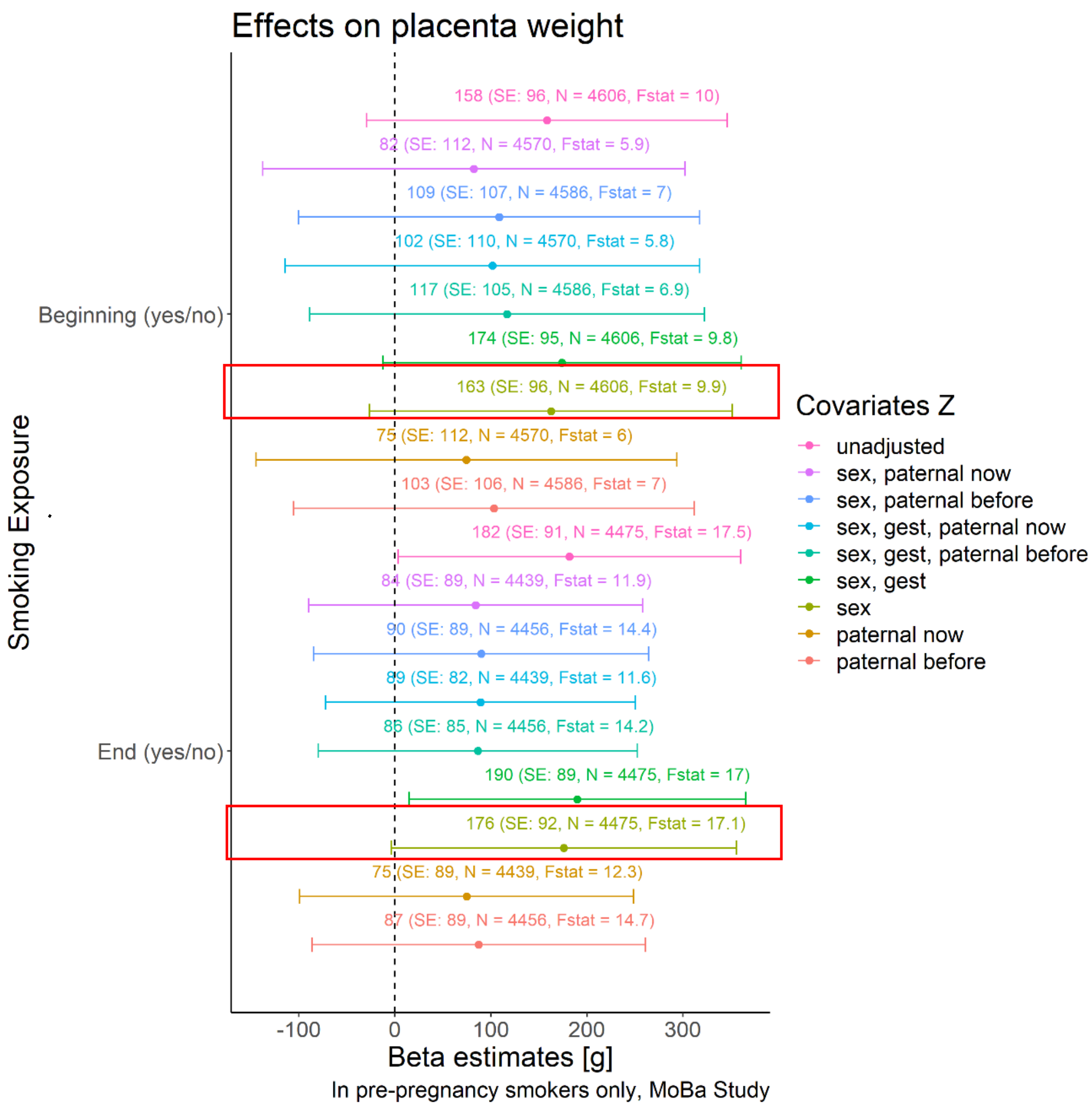


*SFigure 3: Results for the MR analysis of continuing vs quitting smoking in pregnancy while adjusting for different potential confounders of the relationship between the smoking exposure and placental weight. We adjusted for different combinations of the covariates: offspring sex, gestational duration, paternal smoking before pregnancy and paternal smoking at 15 weeks of gestation. In addition to the listed covariates in the legend, all analyses were adjusted for the first 10 principal components of ancestry and the genetic batch variables. The red box highlights the result presented in the main part of the paper.*
